# Detection of SARS-CoV-2 from raw patient samples by coupled high temperature reverse transcription and amplification

**DOI:** 10.1101/2020.05.19.20103150

**Authors:** Johannes W. P. Kuiper, Timo Baade, Marcel Kremer, Ramon Kranaster, Linda Irmisch, Marcus Schuchmann, Johannes Zander, Andreas Marx, Christof R. Hauck

## Abstract

The SARS-CoV-2 beta coronavirus is spreading globally with unprecedented consequences for modern societies. The early detection of infected individuals is a prerequisite for all strategies aiming to contain the virus. Currently, purification of RNA from patient samples followed by RT-PCR is the gold standard to assess the presence of this single-strand RNA virus. However, these procedures are time consuming, require continuous supply of specialized reagents, and are prohibitively expensive in resource-poor settings. Here, we report an improved nucleic-acid-based approach to detect SARS-CoV-2, which alleviates the need to purify RNA, reduces handling steps, minimizes costs, and allows evaluation by non-specialized equipment. The use of unprocessed swap samples and the ability to detect as little as three viral genome equivalents is enabled by employing a heat-stable RNA- and DNA-dependent DNA polymerase, which performs the double task of stringent reverse transcription of RNA at elevated temperatures as well as PCR amplification of a SARS-CoV-2 specific target gene. As results are obtained within 2 hours and can be read-out by a hand-held LED-screen, this novel protocol will be of particular importance for large-scale virus surveillance in economically constrained settings.

## Introduction

As of April 2020, severe acute respiratory syndrome coronavirus 2 (SARS-CoV-2) is responsible for more than 4.1 million COVID-19 cases and associated with more than 285.000 fatalities (WHO; COVID situation report 114 [Accessed 14. May 2020]. Available from: https://www.who.int/emergencies/diseases/novel-coronavirus-2019/situation-reports) Within a few months of its emergence, this infectious agent has spread globally and is derailing societies worldwide. SARS-CoV-2 is an enveloped plus-strand RNA virus of the beta-coronavirus genus with closely related strains circulating in bats and some other mammals indicating a zoonotic origin (Lu, Zhao et al., 2020, Zhou, Yang et al., 2020). Upon host cell infection, beta-coronaviruses generate a minus-strand RNA copy of their genome by a virus-encoded RNA-dependent RNA-polymerase (Snijder, Decroly et al., 2016). Furthermore, the virus directs the synthesis of several subgenomic minus strand RNAs, which are complementary to the 3’ of the viral genome and which encode several non-structural proteins including the N gene (Snijder et al., 2016). Therefore, infected cells harbor plus and minus strand RNAs of the coronavirus, with an overabundance of transcripts derived from the 3’ end of its genome (Den Boon, Spaan et al., 1995, Nedialkova, Gorbalenya et al., 2010). As with other RNA viruses, confirmation of SARS-CoV-2 infection is based on the molecular biological detection of the viral genome and its transcripts in patient samples by nucleic acid amplification techniques (NAATs) (Zumla, Al-Tawfiq et al., 2014). To allow sensitive and accurate detection of viral ribonucleic acids, primary patient samples such as nasopharyngeal swabs, sputum or stool are further processed to isolate the total RNA. Using reverse transcriptase, the RNA is then reverse transcribed into DNA. Next, the DNA is PCR-amplified by a thermo-stable DNA-dependent DNA polymerase using specific primers and probes to detect the presence of SARS-CoV-2 sequences.

Facilitated by genome sequencing and rapid information sharing, such a NAAT-based diagnostic surveillance of SARS-CoV-2 has been in place right from the beginning of the COVID-19 pandemic (Wu, Zhao et al., 2020; Corman, Landt et al., 2020). However, the current methodology is laborious, requires multiple handling steps and expensive consumables, resulting in a costly diagnostic procedure, which can constrain COVID-19 testing in economically tight settings. Furthermore, the overwhelming worldwide demand for some of the same reagents (such as those needed for RNA isolation) and diagnostic kits has produced shortages and unnecessarily delayed or restricted testing. From a clinical point of view, rapid testing and early decision making on further isolation measures for patients and health care workers remains a critical issue.

We have developed a thermostable DNA polymerase, which can mediate DNA synthesis from both RNA as well as DNA templates (Sauter & Marx, 2006). By targeted modifications, we have further improved the accuracy and processivity of this enzyme (Blatter, Bergen et al., 2013), which lays the foundation of the commercialized Volcano3G formulations. We reasoned that such a bi-functional enzyme might allow us to improve and simplify the detection of RNA viruses. Here we show that Volcano3G can be used in a coupled high-temperature reverse transcription and amplification reaction to detect SARS-CoV-2 with high sensitivity and specificity. Moreover, our findings demonstrate the usefulness of such a thermo-stable RNA- and DNA-reading DNA polymerase to simplify COVID-19 diagnostics. Most importantly, we can show that this robust enzyme allows detection of SARS-CoV-2 directly from unprocessed patient material. Accordingly, this streamlined procedure, which does not depend on limited reagents, nor requires expensive equipment, is poised to enable large scale SARS-CoV-2 surveillance in a multitude of resource- or time-constrained settings.

## Results and Discussion

### A RNA- and DNA-reading heat-stable polymerase reverse transcribes and amplifies viral RNA

Evidence of an acute SARS-CoV-2 infection depends on the detection of viral RNA species in patient samples, which necessitates reverse transcription of RNA followed by PCR amplification of the resulting DNA. To investigate, if the Volcano polymerase is able to perform both of these steps, we employed an *in vitro* transcribed synthetic SARS-CoV-2 RNA template covering a ∼750 nt stretch within the 3’-end of the viral genome (Fig. 1A). This target region is also included in the CDC panel of primers (Division of Viral Diseases, National Center for Immunization and Respiratory Diseases, Centers for Disease Control and Prevention, Atlanta, GA, USA; https://www.cdc.gov/coronavirus/2019-ncov/lab/rt-pcr-panel-primer-probes.html; accessed on 5.5.2020). When 5000 genome equivalents of the purified, in vitro transcribed viral RNA was used as a PCR template for a generic, heat-stable DNA-dependent DNA polymerase (Taq DNA polymerase) no amplification occurred, demonstrating the absence of a contaminating DNA template (Fig. 1B). However, upon addition of the Volcano3G polymerase to the mix, the expected amplification product was obtained, confirming that the Volcano3G polymerase can read the RNA template to produce and amplify a specific DNA sequence (Fig. 1B). Clearly, the Taq DNA polymerase was able to yield an amplicon, when DNA instead of RNA was used as a template (Fig. 1B). Not surprisingly, the Volcano3G polymerase was able to operate with primers and probes supplied by other manufacturers targeting the same region of the SARS-CoV-2 N gene (Suppl. Fig. S1A and S1B). Due to the thermostability of the Volcano3G polymerase, this RNA-reading enzyme could perform, in contrast to other widely used enzymes, the reverse transcription under stringent, high-temperature conditions. Reverse transcription at elevated temperatures could help to overcome stable RNA-secondary structures present in beta-coronavirus genomes (Brierley, Digard et al., 1989, Plant, Perez-Alvarado et al., 2005). To take advantage of this particular feature of the Volcano3G polymerase, we added an extended, high-melting temperature virus-specific primer (R2) to the reaction, and adjusted the PCR protocol to include a high-temperature reverse transcription step. As the 3’-end of the viral genomic plus-strand RNA and its minus-strand complementary sequences are the most abundant nucleic acids in coronavirus-infected cells, we focussed on the coronavirus N gene, which is located at the very 3’-end of the SARS-CoV-2 genome (Irigoyen, Firth et al., 2016). Furthermore, the amino terminus of the N protein varies greatly between different human-pathogenic beta-coronavirus isolates (Suppl. Fig. S1C), making this region an ideal target for SARS-CoV-2-specific primers. Accordingly, the high-temperature RT primer R2 was designed to be complementary to sequences at the 5’ end of the N gene (Fig. 1A). The addition of the R2 primer consistently increased the performance of the Volcano3G reaction resulting in lower cq-values over a wide range of template concentrations (Fig. 1C). The high-temperature reverse transcription afforded by Volcano3G polymerase and the R2 primer was optimal at 75°C (Suppl. Fig. S1D). Most importantly, addition of R2 lowered the limit of detection (LOD) to five copies of the viral genome equivalent (Fig. 1D). To assess, if this adapted procedure also allows the sensitive detection of SARS-CoV-2 in patient material, we used RNA isolated from nasopharyngeal swabs of two confirmed COVID-19 cases. In both samples, amplification of the human RNAseP transcript demonstrated the integrity of the samples and resulted in similar cq-values in the presence or absence of the additional R2 primer, while the non-template control (NTC) gave no signal (Fig. 1E). Most importantly, the Volcano3G polymerase detected viral RNA in both samples, but produced exceptionally low cq-values upon addition of the R2 primer, suggesting that the high-temperature reverse transcription by the thermo-stable RNA-reading DNA polymerase opens an additional window of opportunity for hypersensitive detection of viral RNA. Together, these initial results encouraged the further exploration of the Volcano3G polymerase for facilitating the detection of SARS-CoV-2 RNA.

**Figure 1.**
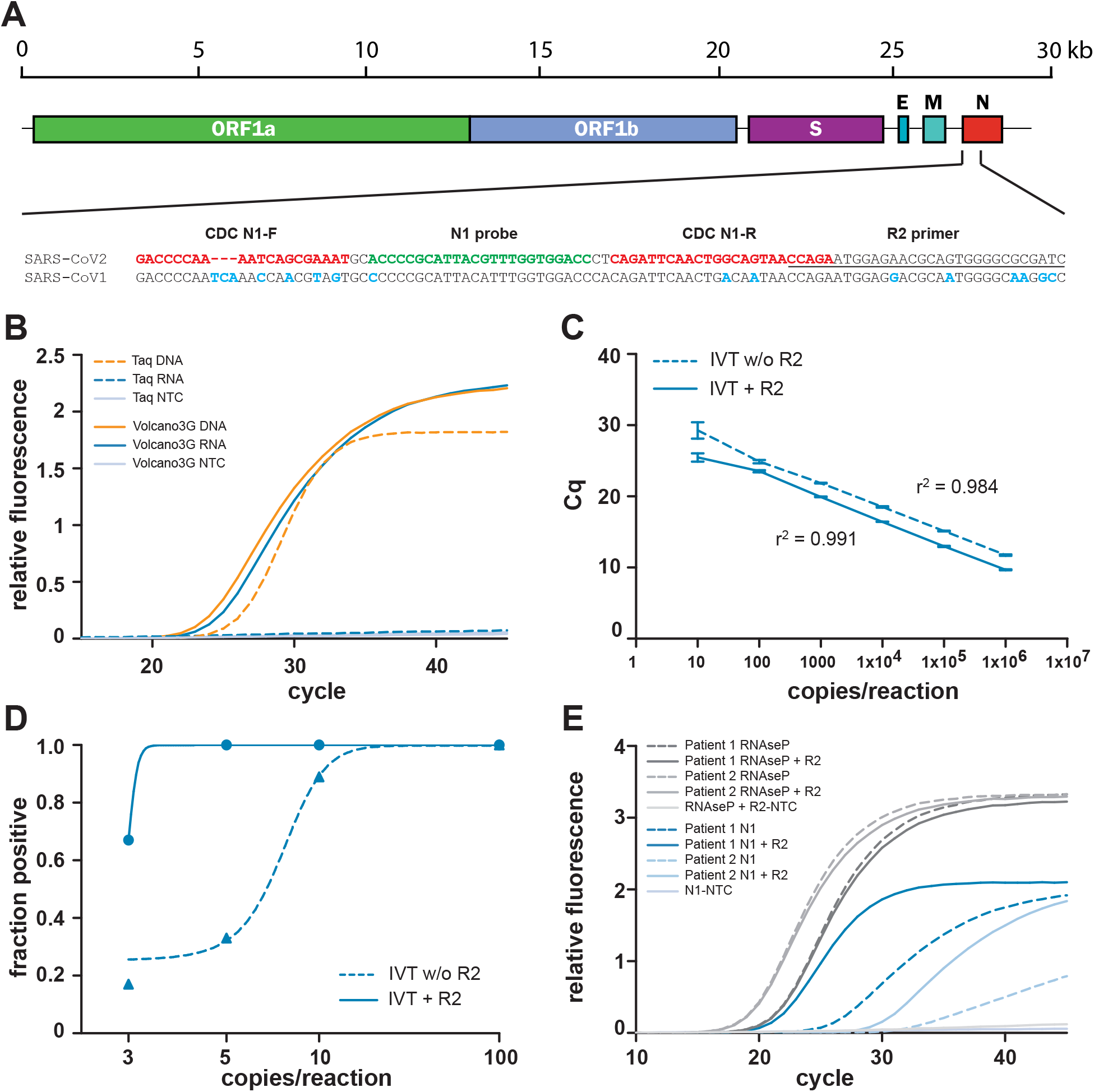
A RNA- and DNA-reading heat-stable polymerase reverse transcribes and amplifies viral RNA. A) Schematic overview of the SARS-CoV-2 genome. The target sequences for the N1 primers and probe are marked in red and green, respectively. The R2 primer binding sequence is underlined. Sequences divergence between SARS-CoV-2 and SARS-CoV-1 genomes are highlighted in blue. **B)** Performance of Volcano3G polymerase was compared to Taq polymerase using plasmid DNA or in vitro transcribed RNA as template (5000 viral genome equivalents). **C)** Determination of the linear dynamic range for the Volcano3G protocol with or without an additional primer (R2) for optimized reverse transcription at a final concentration of 250 nM. In vitro transcribed RNA containing the Sars-CoV-2 N amplicon was serially diluted in the range from 1×10^6^ copies to 10 copies. **D)** Limit of detection (LOD) was assessed with serial dilutions ranging from 20 to 1 copy per reaction (n = 6 for each dilution). The fraction of positive reactions (y-axis) were plotted against the log-transformed number of RNA copies per reaction. Addition of R2 primer enhances the performance at lower copy-numbers. **E)** Amplification curves showing the performance of Volcano3G on isolated RNA from two COVID-19 patients in presence or absence of R2.

### SARS-CoV-2 detection by high-temperature RT-PCR in a patient cohort delivers results consistent with the standard procedure

To evaluate the potential of the high-temperature RT-PCR protocol using Volvano3G for the detection of viral RNAs in patient material, we assessed the presence of SARS-CoV-2 in RNA isolated from a small cohort of COVID-19 suspected cases. RNA was isolated from nasopharyngeal swabs and the isolated nucleic acid was then evaluated in parallel by i) a commercial in vitro diagnostic kit (Allplex, Seegene) and ii) high-temperature RTPCR with Volcano3G. Of the 43 samples, the Allplex-assay detected the SARS-CoV-2 N gene in 35 samples, while 8 samples remained negative (Fig. 2A). The eight negative samples also did not yield amplicons in the Allplex-assay for the RdRp or the E gene (data not shown). When the 43 RNA samples were employed in high-temperature RT-PCR with Volcano3G polymerase, the identical results were obtained, showing complete consistency with regard to positive and negative outcomes (Fig. 2A and 2B). Pairwise comparison of the cq-values obtained for the isolated RNA samples in detecting the N gene revealed that the high-temperature RT-PCR with Volcano3G resulted in lower cq-values throughout all samples suggesting a slightly increased sensitivity (Fig. 2C). In addition to the resolution of RNA secondary structures, the increased sensitivity might be due to the fact that the high temperature reverse transcription step involves several cycles, which allow initial highly stringent amplification of viral target genes. Though the high temperature RT-PCR with Volcano3G was slightly more sensitive, the results correlated extremely well over a wide range of cq-values with the results obtained by the commercial assay (r^2^ = 0.980, p<0.0001, Fig. 2D). Together, these findings suggest that high temperature RT-PCR with Volcano3G could be an additional option to rapidly detect RNA-virus genomes with high specificity and sensitivity.

**Figure 2.**
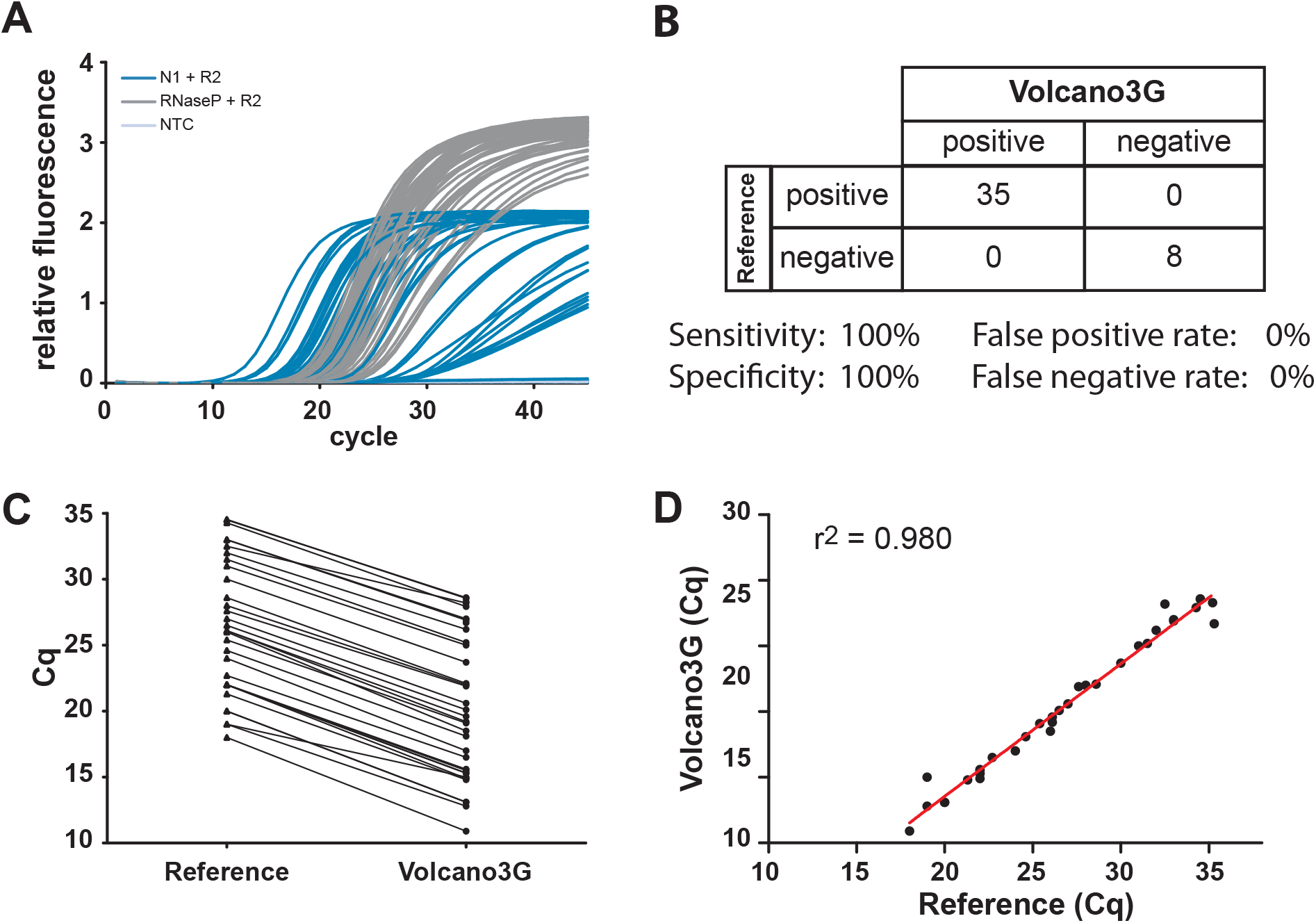
SARS-CoV-2 detection by high-temperature RT-PCR in a patient cohort delivers results consistent with the standard procedure. **A**) RNA was isolated from nasopharyngeal- and throat swab samples (n = 43) and SARS-CoV-2 and RNAseP were detected using the Volcano3G protocol. N1 amplicon (blue), RNaseP gene (gray). Water was used as a non-template control (light gray). **B)** Identical samples were processed in parallel in an accredited diagnostic lab using the Allplex 2019-nCoV assay from Seegene. Direct comparison of assay results reveals 100% concordance of Volcano3G with the reference assay. **C)** Cq values obtained with Volcano3G were lower than those obtained with the reference assay (ΔCq = 6.4 +/− 0.78). **D)** For each positive patient sample, the Cq values obtained with both assays were plotted against each other for linear regression analysis. A highly significant correlation of Volcano3G with the reference assay was observed (r^2^ = 0.98, p<0.0001).

### High-temperature RT-PCR using Volcano3G polymerase allows SARS-CoV-2 detection from unprocessed patient samples

Besides enhanced stringency and possible resolution of RNA secondary structures, we speculated that high-temperature RT-PCR might promote viral lysis and allow the detection of SARS-CoV-2 RNA directly from unprocessed patient samples. To this end, we employed a second cohort of samples, where each nasopharyngeal swab was initially resolved in 300 µl of distilled water. While 150 µl of these diluted samples were used for RNA extraction and standard RT-PCR, 12 µl of the diluted sample were directly employed in high-temperature RT-PCR with the Volcano3G polymerase. Importantly, even with this raw patient material, the high-temperature RT-PCR yielded clear-cut results without increasing the background (Fig. 3A). When directly compared to the results obtained by the standard RT-PCR from purified RNA of the identical samples, all negative samples were consistent between the two approaches suggesting that there is no increased risk of producing false positive results when using the unprocessed patient samples (Fig. 3B). Moreover, 100% of the samples showing cq-values of ≤ 30 for the SARS-CoV-2 N gene in the standard RT-PCR assay with purified RNA were correctly identified by the Volcano3G high-temperature RT-PCR employing raw patient samples (Fig. 3B). Samples yielding cq-values above 30 in the standard RT-PCR following RNA extraction, which would represent a low viral load, were only rarely detected as positive, when patient samples were used without further processing (1 in 5 samples, Fig. 3B). None of these 5 patients with low viral loads was hospitalized and all of them showed only mild symptoms including sore throat and rhinitis. It is important to stress that clinical studies of viral dynamics in patients have shown that SARS-CoV-2 titers in swab samples are most often high during the initial phase of the infection, even before onset of symptoms (Pan, Zhang et al., 2020, Zou, Ruan et al., 2020). In contrast, low titers are typically seen at the later phase of the infection and can even be detected after the resolution of symptoms (Lescure, Bouadma et al., 2020). If infectious virus is spread in situations, where viral titers are low, is still unclear.

**Figure 3.**
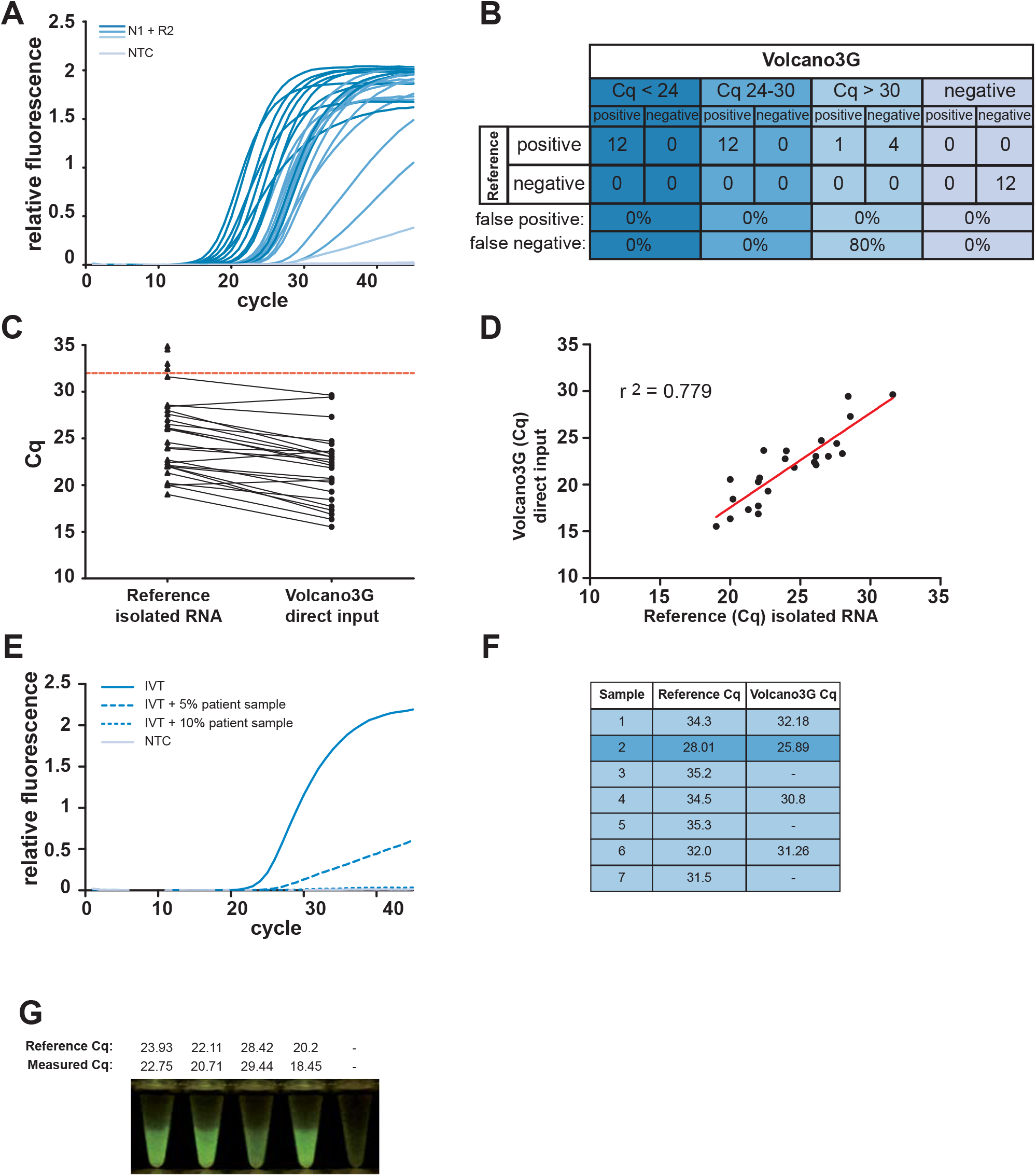
High-temperature RT-PCR using Volcano3G polymerase allows SARSCoV-2 detection from unprocessed patient samples. **A)** Nasopharyngeal- and throat swab samples (prepared in water) were added directly as template for RT-qPCR using the Volcano3G protocol. Representative amplification curves of patients with high (dark blue), medium (medium blue) and low Cq as well as negative patients (light blue) are shown. **B)** RNA was isolated from the remaining patient material and analysed in an accredited diagnostic lab using the Allplex 2019-nCoV assay from Seegene. **C)** The Cq values of each patient sample are compared between the reference protocol and the Volcano3G direct approach. Dotted red line indicates the cut-off, were the assay loses sensitivity. **D)** For each positive patient sample, the Cq values obtained with both assays were plotted against each other for linear regression analysis (r^2^ = 0.779, p<0.0001) **E)** RTPCR analysis of 100 copies of in vitro transcribed RNA spiked with varying amounts of pooled patient material from 5 confirmed negative patients. **F)** RT-PCR analysis of confirmed COVID-19 patient samples with high Cq values were analysed in a larger volume. **G)** Four confirmed COVID-19 patient samples and one negative control were used directly as in A). The reference cq-values obtained by standard RT-PCR from purified RNA (upper row) and the cq-values obtained by high temperature RT-PCR with Volcano3G polymerase (loer row) are given. After completion of PCR cycling, the reaction tubes were photographed on a blue light transilluminator (470 nm). Positive samples emitted a distinct green fluorescence visible by the naked eye.

Interestingly, for most positive samples detected by the high-temperature RT-PCR with Volcano3G, the cq-values were lower compared to the standard RT-PCR (Fig. 3C and D), indicating that the detection of SARS-CoV-2 from unprocessed patient material might not be limited by the sensitivity of this direct approach. As unprocessed swab material might contain numerous patient-derived proteins, mucus, or membrane fragments, we speculated that substance(s) within these raw samples could interfere and inhibit the detection of SARS-CoV-2 when only a low viral load is present. Therefore, we tested the inhibitory potential of the eluted swab material. Using the in vitro transcribed viral RNA as a template, we spiked the Volcano3G reaction mix with increasing amounts of swab-derived material (Fig. 3E). Importantly, there was a clear dose-dependent inhibition of the amplification reaction due to the material eluted from the swabs (Fig. 3E and Suppl. Fig. S2). The extent of inhibition showed variability between individual samples (Suppl. Fig. S2), with some samples leading to complete inhibition of viral detection at 10% added swab material (Fig. 3E). Especially, for samples containing low viral titers this could significantly elevate the LOD. Therefore, we supplemented the distilled water used for eluting the nasopharyngeal swabs with betaine, BSA, carrier RNA, DTT or treated them with proteinase K or combinations of these reagents, with the idea that these substances might be able to alleviate the inhibition due to inhibitory proteins or RNA degrading or oxidizing agents. Indeed, several of the treatments enhanced the detection of SARS-CoV-2 from an unprocessed patient sample demonstrating that it is possible to partially overcome the inhibitory effect of the raw material (Suppl. Fig. 3A and B). Furthermore, we reasoned that increasing the volume of the high-temperature RT-PCR reaction to allow an increase of viral copies without increasing the relative abundance of inhibitory factors could also help in improving the detection of SARS-CoV-2 in low-titer patient samples. Using addition of carrier RNA and DTT as well as an increased input, we assayed an additional set of five SARS-CoV-2 positive samples, which had produced ct values > 30 for the N gene in the standard RT-PCR procedure with purified RNA (Fig. 3F). Interestingly, 3 of the 5 samples could now be detected as positive, when the unprocessed samples were used as input for the high temperature RT-PCR with Volcano3G polymerase (Fig. 3F). These results demonstrate that even low virus titers can be detected directly from raw patient samples by employing the high temperature RT-PCR conditions. Though further improvements will be necessary to consistently detect SARS-CoV-2 RNA in unprocessed low-titer samples, bypassing the need for RNA purification dramatically accelerates and simplifies the evaluation of patient material. As an additional feature, we have observed that the endpoint of the high temperature RT-PCR with Volcano3G polymerase can also be easily evaluated on a blue-light LED screen, yielding strong green fluorescence for positive samples and allowing clear-cut differentiation by the naked eye (Fig. 3G).

Together, the use of a heat-stable, RNA- and DNA-reading polymerases is poised to simplify detection of viral RNAs. By reducing handling steps, the chances of sample cross-contamination are minimized. The overwhelming dynamic spread of SARS-CoV-2 and the apparent bottle-necks in virus testing highlight the need for additional methodology that can be deployed in resource poor settings and that is operative in situations, where particular reagents such as RNA isolation kits might become scarce. Therefore, the direct detection of virus RNA from unprocessed patient samples and the ability to perform and read out complex NAAT procedures using standard field PCR machines and blue-light screens could dramatically expand the COVID-19 testing capabilities and might also afford an important economic relief for health care providers in large parts of the world.

## Material and Methods

### Patient samples and ethics statement

Nasopharyngeal swabs were collected at the COVID-19 test center hosted at the Klinikum Konstanz. Samples used in this study were procured with sterile dry swabs (Copan 160C Rayon, http://www.copaninnovation.com; Eurotubo collection swab 300253, http://www.deltalab.es; culture swab without transport medium 09–516–5009, http://www.nerbe-plus.de) and samples were processed within 24h. Patients gave informed consent to participate in the study “*Entnahme von Schleimhautabstrichen zur Detektion von Sars-CoV-2 bei Verdacht auf COVID-19*” registered at the German Clinical trials Register (DRKS00021578). The study was approved by the local ethics committee of University of Konstanz (05/2020).

### RNA extraction

Nasopharyngeal swabs were eluted in 350 µl RNAse/DNAse free deionized water and 150 μl of the eluate was used for RNA isolation using the NucleoSpin 96 RNA kit (Macherey-Nagel, Düren, Germany), where the RA1 lysis buffer included in the kit was supplemented with 10 µg/ml of carrier RNA (Qiagen, Hilden, Germany) and 15 mM DTT. Automated handling of the samples was accomplished using the Integra Assist Plus pipetting robot (Integra, Biebertal, Germany). Finally, purified RNA was eluted into 60 µl RNAse/DNAse free deionized water.

### Generation of in vitro transcribed N1 amplicon

cDNA was generated from isolated viral RNA using the Maxima cDNA synthesis kit (Fermentas, Vilnius, Lithuania) according to the manufacturer’s protocol. A region corresponding to position 27923–28648 of the SARS-CoV2 genome (NC_045512.2) was amplified with Pfu DNA polymerase (primers Clone_N1_F and Clone_N1_R, see table 1). The resulting 726 bp amplicon was purified, digested (KpnI/BamHI) and ligated in pBluescript-KS(+). The resulting plasmid was linearized with BamHI, purified and transcribed in vitro (T7 DNA polymerase, NEB) for 4 hours at 37°C to yield a 753 nt transcript. The purified transcript was treated with RNAse-free DNase (Roche) for 4 hours at 37°C and purified again (viral RNA isolation kit, Qiagen).

**Table 1:**
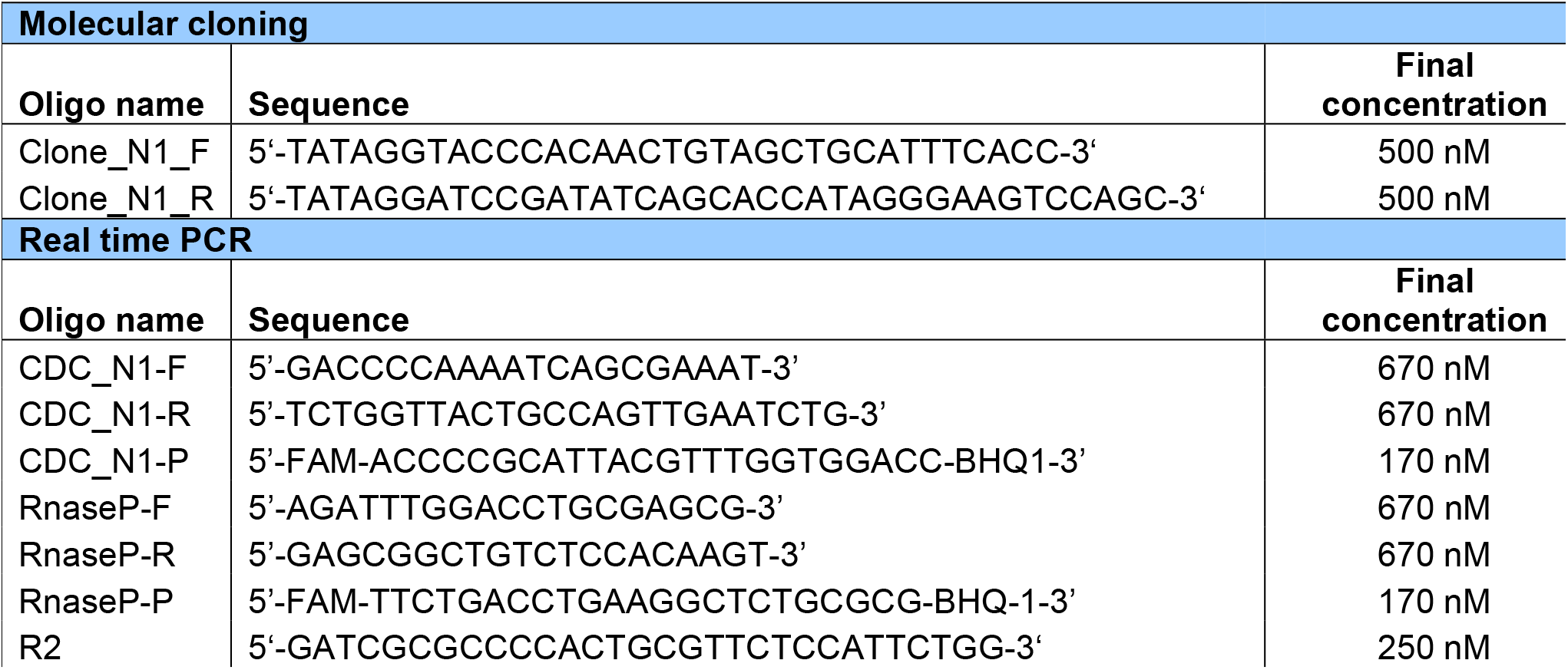
Primer and probe sequences used in this study

### Standard RT-PCR with purified RNA

Reference Cq values were obtained using the Allplex 2019-nCoV Assay (Seegene, Seoul, South Korea) according to the manufacturer’s instructions. 8 μL of purified RNA were used to run 25 µl reactions on a Bio-Rad CFX96 and analysed with Seegene Viewer Software version 3.18.

### High temperature RT-PCR with purified RNA

Throughout this study high-temperature RT-PCR on RNA purified from either the in vitro transcribed viral genome fragment or the swab material was performed with the Volcano3G RT-PCR Probe 2x Master Mix (in short: Volcano3G) (myPOLS Biotec, Konstanz, Germany) using the CDC-approved N1 primer/probe mix from Integrated DNA Technologies (IDT, San Diego, CA, USA). In addition, sequence-identical primers and probe were obtained from Microsynth AG (Balgach, Switzerland) to produce data presented in Supplemental Fig. S1. Typically, 10 µl reactions were set up containing 1 µl RNA, 1x Volcano3G MasterMix, CDC_N1 or RNaseP primer/probe mix (see table 1) and 250 nM R2 primer. A three-stage thermal cycling program was performed on a Roche LightCycler 96, consisting of 1) 150 seconds at 75°C, 2) 10 cycles of 95°C for 5 seconds, 150 seconds at 75°C, 3) 45 cycles of 95°C for 10 seconds, 57°C for 35 seconds. All assays were performed in white low profile tubes with ultra-clear caps (ThermoFisher, Cat No: AB1771) as singleplex with FAM/BHQ1-labelled probes. Cq values were determined with Roche LightCycler Software 1.1.0.

### High temperature RT-PCR with unprocessed patient material

Nasopharyngeal swabs were eluted in 350 µl RNAse/DNAse free deionized water. These unprocessed samples were added directly to the PCR reaction. Reaction conditions were identical to those described for purified RNA as template, with the exception that the reaction volume was increased to 50 µl and 12 µl unprocessed sample was used (unless otherwise stated in the results section). In addition, successful amplification could be visualised by detection of the dequenched fluorescein signal (FAM) on a blue light transilluminator (Safe Imager 2.0, Invitrogen).

### Treatments for improving high-temperature RT-PCR of raw patient samples

In an effort to improve the efficiency of our protocol on unprocessed patient samples, several treatment regimens were tested. Nasopharyngeal swab samples in distilled water were supplemented with 1 ng/µl carrier RNA (Jena Analytik), 100 mM betaine (molecular grade, Sigma Aldrich), 0.05% bovine serum albumin (BSA) or a combination of all three, before adding the samples directly to the Volcano3G master mix. In another approach, swab samples were treated with 128 µg/µl nuclease-free proteinase K (Analytik Jena) for 10 minutes at 70°C in 5 mM HEPES (pH 7.4) followed by inactivation for 10 minutes at 95°C. Alternatively, samples were treated with 2.5 mM nuclease-free DTT (Thermo Fisher Scientific) for 10 minutes at 70°C or a combination of proteinase K and DTT.

### Limit of detection (LOD) analysis

Serial dilutions of the in vitro transcribed amplicon were made in water containing 5 ng/µl RNA carrier mix (innuPREP Virus DNA/RNA Kit, Analytik Jena) and real-time PCR reactions (10 µl reaction volume) were set up as described above. Three to eight replicate reactions were conducted containing 1×10^6^, 1× 10^5^, 1×10^4^, 1×10^3^, 100, 10, 5, 3 or 1 copies per reaction. To estimate the LOD, the fraction of positive reactions (Cq < 40) was plotted against the log-transformed number of amplicon copies.

### Statistical analysis

Linear regression analysis was performed with Prism (GraphPad, La Jolla, CA, USA).

## Data Availability

All original data are included in the manuscript

## Acknowledgements

We thank the members of the AG Hauck and AG Groettrup for their help in processing patient samples, Thomas Meyer and Silke Müller from the Screening Center of University of Konstanz for automated RNA isolation, and Susanne Feindler-Boeckh for expert technical assistance. TB acknowledges support from the Konstanz Research School Chemical Biology (KoRS-CB) and AM support by the German Research Foundation (DFG) within SPP 1784.

## Author contributions

JK, TB: conceived and performed experiments, analysed data, prepared figures and drafted the manuscript; MK, JZ: analysed samples and data, commented on the manuscript; LI, MS: recruited patients, procured samples, commented on the manuscript; RK, AM: provided essential reagents, consulted on experiments and commented on the manuscript; CRH: conceived the study, analysed data, wrote the manuscript.

---

**Supplementary Figure S1. The R2 reverse primer enhances detection of the viral N gene RNA. A)** N1 oligonucleotides and fluorescent probes according to CDC’s recommendations were ordered from an alternative manufacturer (Microsynth AG) and assessed for their performance using Volcano3G and 500 copies of in vitro transcribed RNA encoding the N1 amplicon. Concentration of forward and reverse primers were adjusted for optimal performance. **B)** The chosen primer/probe concentrations were evaluated using isolated RNA from two confirmed SARS-CoV-2 positive patients, using RNAseP as control (dashed grey lines). Dashed blue lines: primer/probe pair without R2. Addition of R2 reverse primer at a final concentration of 250 nM enhanced the performance of the N1 primer pair, while showing no effect on RNAseP amplification (solid blue and grey lines, respectively). Light grey line: non-template control. **C)** Amino acid sequences of the amino-terminus of the beta-coronavirus N gene from SARS-CoV-2, SARS-CoV-1, MERS, and the human coronavirus strains OC43, HKU1, 229E, and NL63. Identical amino acids are marked in red. The N protein displays high sequence divergence at the amino terminus suggesting that the corresponding region of the N gene (upper row) is well suited for selective detection of SARS-CoV-2. **D)** Isolated RNA from a confirmed SARS-CoV-2 positive patient was analysed using the Volcano3G protocol and the N1 primer/probe mix from IDT in presence of 250 nM of R2. A temperature gradient was run during the reverse transcription reaction (step 1 and 2) of the Volcano3G thermocycling program.

**Supplementary Figure S2. Swab-derived material contains inhibitory factors. A)** Nasopharyngeal swab samples from three confirmed negative patients were serially diluted in RNAse free water and spiked with 5000 copies of in vitro transcribed RNA revealing the presence of PCR inhibitors. **B)** Unprocessed patient material from two confirmed SARS-CoV2 positive patients and isolated RNA from one confirmed positive patient were serially diluted in RNAse free water containing carrier RNA (1 ng/µl). Empirically determined Cq values are plotted against a theoretical dilution curve.

**Supplementary Figure S3. The inhibitory effects of raw patient material can be ameliorated. A)** An unprocessed nasopharyngeal swab sample of a confirmed SARSCoV-2 positive patient was diluted in water plus either carrier RNA (1ng/µl), betaine (100 mM), BSA (0.05%) or a combination of all three and subjected to Volcano RT-qPCR. **B)** Nasopharyngeal swab sample of a confirmed positive patient was diluted in water plus carrier RNA (1 ng/µl). Patient material was then treated with Proteinase K (ProtK, 128 µg/ml), DTT (2.5 mM) or a combination of both. Samples were incubated at 70°C for 10 min. Samples containing ProtK were additionally inactivated at 95°C for 10 min.

## References

Blatter N, Bergen K, Nolte O, Welte W, Diederichs K, Mayer J, Wieland M, Marx A (2013) Structure and function of an RNA-reading thermostable DNA polymerase. Angew Chem Int Ed Engl 52: 11935–9

Brierley I, Digard P, Inglis SC (1989) Characterization of an efficient coronavirus ribosomal frameshifting signal: requirement for an RNA pseudoknot. Cell 57: 537–47

Corman VM, Landt O, Kaiser M, Molenkamp R, Meijer A, Chu DK, Bleicker T, Brunink S, Schneider J, Schmidt ML, Mulders DG, Haagmans BL, van der Veer B, van den Brink S, Wijsman L, Goderski G, Romette JL, Ellis J, Zambon M, Peiris M et al. (2020) Detection of 2019 novel coronavirus (2019-nCoV) by real-time RT-PCR. Euro Surveill 25: pii = 2000045

Den Boon JA, Spaan WJ, Snijder EJ (1995) Equine arteritis virus subgenomic RNA transcription: UV inactivation and translation inhibition studies. Virology 213: 364–72

Irigoyen N, Firth AE, Jones JD, Chung BY, Siddell SG, Brierley I (2016) High-Resolution Analysis of Coronavirus Gene Expression by RNA Sequencing and Ribosome Profiling. PLoS Pathogens 12: e1005473

Lescure FX, Bouadma L, Nguyen D, Parisey M, Wicky PH, Behillil S, Gaymard A, Bouscambert-Duchamp M, Donati F, Le Hingrat Q, Enouf V, Houhou-Fidouh N, Valette M, Mailles A, Lucet JC, Mentre F, Duval X, Descamps D, Malvy D, Timsit JF et al. (2020) Clinical and virological data of the first cases of COVID-19 in Europe: a case series. The Lancet Infectious Diseases doi: 10.1016/S1473-3099(20)30200-0. [Epub ahead of print]

Lu R, Zhao X, Li J, Niu P, Yang B, Wu H, Wang W, Song H, Huang B, Zhu N, Bi Y, Ma X, Zhan F, Wang L, Hu T, Zhou H, Hu Z, Zhou W, Zhao L, Chen J et al. (2020) Genomic characterisation and epidemiology of 2019 novel coronavirus: implications for virus origins and receptor binding. Lancet 395: 565–574

Nedialkova DD, Gorbalenya AE, Snijder EJ (2010) Arterivirus Nsp1 modulates the accumulation of minus-strand templates to control the relative abundance of viral mRNAs. PLoS Pathogens 6: e1000772

Pan Y, Zhang D, Yang P, Poon LLM, Wang Q (2020) Viral load of SARS-CoV-2 in clinical samples. The Lancet Infectious Diseases 20: 411–412

Plant EP, Perez-Alvarado GC, Jacobs JL, Mukhopadhyay B, Hennig M, Dinman JD (2005) A three-stemmed mRNA pseudoknot in the SARS coronavirus frameshift signal. PLoS Biology 3: e172

Sauter KB, Marx A (2006) Evolving thermostable reverse transcriptase activity in a DNA polymerase scaffold. Angew Chem Int Ed Engl 45: 7633–5

Snijder EJ, Decroly E, Ziebuhr J (2016) The Nonstructural Proteins Directing Coronavirus RNA Synthesis and Processing. Advances in Virus Research 96: 59–126

Wu F, Zhao S, Yu B, Chen YM, Wang W, Song ZG, Hu Y, Tao ZW, Tian JH, Pei YY, Yuan ML, Zhang YL, Dai FH, Liu Y, Wang QM, Zheng JJ, Xu L, Holmes EC, Zhang YZ (2020) A new coronavirus associated with human respiratory disease in China. Nature 579: 265–269

Zhou P, Yang XL, Wang XG, Hu B, Zhang L, Zhang W, Si HR, Zhu Y, Li B, Huang CL, Chen HD, Chen J, Luo Y, Guo H, Jiang RD, Liu MQ, Chen Y, Shen XR, Wang X, Zheng XS et al. (2020) A pneumonia outbreak associated with a new coronavirus of probable bat origin. Nature 579: 270–273

Zou L, Ruan F, Huang M, Liang L, Huang H, Hong Z, Yu J, Kang M, Song Y, Xia J, Guo Q, Song T, He J, Yen HL, Peiris M, Wu J (2020) SARS-CoV-2 Viral Load in Upper Respiratory Specimens of Infected Patients. New England Journal of Medicine 382: 1177–1179

Zumla A, Al-Tawfiq JA, Enne VI, Kidd M, Drosten C, Breuer J, Muller MA, Hui D, Maeurer M, Bates M, Mwaba P, Al-Hakeem R, Gray G, Gautret P, Al-Rabeeah AA, Memish ZA, Gant V (2014) Rapid point of care diagnostic tests for viral and bacterial respiratory tract infections--needs, advances, and future prospects. The Lancet Infectious Diseases 14: 1123–1135

